# Association of Clonal Hematopoiesis of Indeterminate Potential with Incident Heart Failure with Preserved Ejection Fraction

**DOI:** 10.1101/2023.06.07.23291038

**Authors:** Alex P. Reiner, Mary B. Roberts, Michael C. Honigberg, Charles Kooperberg, Pinkal Desai, Alexander G. Bick, Pradeep Natarajan, JoAnn E. Manson, Eric A. Whitsel, Charles B. Eaton

**Affiliations:** Public Health Sciences Division, Fred Hutchinson Cancer Center, Seattle, Washington; Center for Primary Care and Prevention, Brown University, Pawtucket, Rhode Island; Broad Institute of Harvard and MIT, Cambridge, Massachusetts; Harvard Medical School, Boston, Massachusetts; Division of Cardiology, Department of Medicine, Massachusetts General Hospital, Boston, Massachusetts; Division of Hematology and Oncology, Weill Cornell Medical College, New York, New York; Division of Genetic Medicine, Department of Medicine, Vanderbilt University School of Medicine, Nashville, Tennessee; Cardiovascular Disease Initiative of the Broad Institute of Harvard and MIT, Cambridge, Massachusetts; Division of Preventive Medicine, Brigham and Women’s Hospital, Boston, Massachusetts; Department of Epidemiology, Gillings School of Global Public Health and Department of Medicine, School of Medicine, University of North Carolina, Chapel Hill, North Carolina; Department of Epidemiology, Brown University, Providence, Rhode Island; Care New England, Center for Primary Care and Prevention, Pawtucket, Rhode Island; Department of Family Medicine, Warren Alpert Medical School of Brown University, Providence, Rhode Island

## Abstract

**Background:** Clonal hematopoiesis of indeterminate potential (CHIP) was recently identified as a risk factor for incident heart failure (HF). Whether CHIP is associated selectively with risk of heart failure with reduced ejection fraction (HFrEF) or heart failure with preserved ejection fraction (HFpEF) subtypes is unknown

**Objectives:** To evaluate whether CHIP is associated with incident HF subtypes, HFrEF versus HFpEF.

**Methods:** We obtained CHIP status from whole genome sequencing of blood DNA in participants without prevalent HF from a multi-ethnic sample of post-menopausal women without prevalent HF (N=5,214) from the Women’s Health Initiative (WHI). Cox proportional hazards models were performed, adjusting for demographic and clinical risk factors.

**Results:** CHIP was significantly associated with a 42% (95%CI 6%, 91%) increased risk of HFpEF (P=0.02). In contrast, there was no evidence of association between CHIP and risk of incident HFrEF. When the three most common CHIP subtypes were assessed individually, the risk of HFpEF was more strongly associated with TET2 (HR=2.5; 95%CI 1.54, 4.06; P<0.001), than DNMT3A or ASXL1.

**Conclusion:** CHIP, particularly mutations in *TET2*, represents a potential new risk factor for incident HFpEF.

## Introduction

Clonal hematopoiesis of indeterminate potential (CHIP) is characterized by the age-related clonal expansion of somatic mutations (most commonly in *DMNT3A, TET2, ASXL1*, and *JAK2*) in hematopoietic cells. It is now well-established that CHIP, as detected in peripheral blood of otherwise healthy individuals, is a risk factor for atherosclerotic cardiovascular disease (CVD) events including coronary heart disease (CHD), stroke, and CVD-related mortality^1^. CHIP also has been associated with adverse outcomes (recurrent hospitalizations and mortality) in patients with pre-existing ischemic heart failure (HF)^2^ and was recently identified as an independent risk factor for incident HF in a meta-analysis of >56,000 individuals from 5 population-based cohorts^3^. Moreover, experimental models of HF have suggested a direct role of CHIP pro-inflammatory myeloid cells on cardiac dysfunction through myocardial fibrotic remodeling and hypertrophy^2^. In observational studies, the association between CHIP and HF has been similar in individuals with and without prior CHD^3^, suggesting the possibility that CHIP may be involved directly in HF pathophysiology rather than simply reflecting the strong link between CHIP and atherosclerosis. Understanding whether CHIP is associated specifically with risk of heart failure with reduced or preserved ejection fraction (HFrEF or HFpEF) may help address this question.

## Methods

We evaluated the association of CHIP with incident HFpEF and HFrEF utilizing data from a multi-ethnic sample of post-menopausal women without prevalent HF from the Women’s Health Initiative (WHI). The Women’s Health Initiative (WHI) recruited 161,808 post-menopausal women aged 50-79 years between 1993 and 1998 at 40 clinical centers in the United States. The WHI participants for the current analysis (N=11,085) were part of a nested case-control sample of incident stroke and venous thromboembolism (VTE), all of whom underwent whole genome sequencing (WGS) at baseline through the NHLBI TOPMed project. CHIP was detected from WGS based on 74 pre-specified driver mutations as previously described^3^. A subset of these WHI TOPMed participants who were randomized to the Hormone Trial and all Black and Hispanic participants (total N=5,214) was followed through March 1, 2020, by yearly medical record abstraction of all self-reported hospitalizations and classified by physician adjudicators as HFrEF or HFpEF^3^. The Institutional Review Board at the Fred Hutchinson Cancer Center approved the studies and participants provided written informed consent.

## Results

At baseline, the mean age of the 5,214 women was 62 years, 70% were White, and 7.8% had CHIP. During a median of 15 years follow-up, 301 (5.8%) developed incident HFpEF and 213 (4.1%) developed HFrEF. The age-adjusted HFpEF incidence rates were 5.86 (95%CI 5.63, 6.10) versus 3.72 (3.68, 3.77) per 1,000 person-years among CHIP carriers versus non-carriers. The age-adjusted HFrEF incidence rates were 2.65 (2.54, 2.76) versus 2.72 (2.68, 2.75) per 1,000 person-years among CHIP carriers versus non-carriers.

In Cox proportional hazards models using inverse-probability weighting to account for the TOPMed sampling scheme and further adjusting for demographic and clinical risk factors (age, race, income, education, smoking status, systolic blood pressure, anti-hypertensive medication use, and history of diabetes, CVD, or atrial fibrillation), CHIP was significantly associated with a 42% (95%CI 6%, 91%) increased risk of HFpEF (P=0.02). In contrast, there was no evidence of association between CHIP and risk of incident HFrEF (**Table 1**). When the three most common CHIP subtypes were assessed individually, the risk of HFpEF was more strongly associated with *TET2* (HR=2.5; 95%CI 1.54, 4.06; P<0.001), than *DNMT3A* or *ASXL1* (**Table 1**).

**Table 1:** Association of CHIP and its Major Subtypes with HFpEF and HFrEF.

| **Outcome** | **Cases/Total N** | **CHIP Exposure** | **Hazard ratio (95% CI)** | **P-value** |
| --- | --- | --- | --- | --- |
| HFpEF* | 301/5001 | Any CHIP | 1.42 (1.06, 1.91) | 0.020 |
|  |  | *DNMT3A* | 1.11 (0.74, 1.67) | 0.624 |
|  |  | *TET2* | 2.50 (1.54, 4.06) | <0.001 |
|  |  | *ASXL1* | 1.04 (0.31, 3.54) | 0.953 |
| HFrEF† | 213/4913 | Any CHIP | 1.02 (0.67, 1.55) | 0.925 |
|  |  | *DNMT3A* | 0.62 (0.32, 1.21) | 0.164 |
|  |  | *TET2* | 0.95 (0.38, 2.36) | 0.911 |
|  |  | *ASXL1* | 2.01 (0.70, 5.79) | 0.194 |

\*HFrEF and HF unknown (HFuEF) were censored at the time of HFpEF. †HFpEF and HFuHF were censored at the time of HFrEF. Cox regression models were adjusted for age, race, income, education, smoking status, systolic blood pressure, anti-hypertensive medication use, and history of diabetes, CVD, or atrial fibrillation. The TOPMed sampling scheme was accounted for using inverse probability weighting.

## Discussion

In an analysis of post-menopausal women free of HF, CHIP (as defined by somatic mutations in leukemia-related genes in the absence of hematologic malignancy) was associated prospectively with increased risk of a first episode of HFpEF, independently of traditional risk factors. In contrast, we did not find evidence for association of CHIP with incident HFrEF. In analyses of specific CHIP driver mutations, *TET2*, was most strongly associated with risk of incident HFpEF.

HFpEF is a heterogeneous clinical syndrome common in older adults. While inflammation represents a common mechanism involved in both HFpEF and HFrEF, chronic, low-grade inflammation and fibrotic remodeling driven by myeloid cells infiltrating the myocardium is increasingly recognized as a primary contributor to HFpEF as well as its aging-related comorbidities^5^. Previous animal models of CHIP, particularly the *TET2* subtype, support the role of CHIP-induced inflammatory cytokine production and inflammatory cell signaling in the pathogenesis of cardiac remodeling, hypertrophy, fibrosis, and ventricular dysfunction induced by either myocardial ischemia or pressure-overload^2^. Our finding that CHIP was specifically associated with increased risk of HFpEF further supports the notion that CHIP may contribute directly to non-ischemic forms of HF, independently of atherosclerosis. If confirmed in additional human observational studies, further elucidation of the connection between CHIP, inflammatory signaling pathways, and HFpEF may have potential therapeutic implications for future management of HFpEF including use of targeted anti-inflammatory therapies such IL-1beta or NLRP3 inflammasome inhibitors^5^.

## Data Availability

All data produced in the present study are available upon reasonable request to the authors.

## Funding

Whole genome sequencing (WGS) for the Trans-Omics in Precision Medicine (TOPMed) program was supported by the National Heart, Lung and Blood Institute (NHLBI). WGS for “NHLBI TOPMed: Trans-Omics for Precision Medicine Whole Genome Sequencing Project: WGS for “NHLBI TOPMed: Women’s Health Initiative (WHI)” (phs001237) was performed at the Broad Institute (HHSN268201500014C). Centralized read mapping and genotype calling along with variant quality metrics and filtering were provided by the TOPMed Informatics Research Center (3R01HL-117626-02S1; contract HHSN268201800002I). Phenotype harmonization, data management, sample-identity QC, and general study coordination, were provided by the TOPMed Data Coordinating Center (3R01HL-120393-02S1; contract HHSN268201800001I). We gratefully acknowledge the studies and participants who provided biological samples and data for TOPMed.

The WHI program is funded by the National Heart, Lung, and Blood Institute, National Institutes of Health, U.S. Department of Health and Human Services through contracts HHSN268201600018C, HHSN268201600001C, HHSN268201600002C, HHSN268201600003C, and HHSN268201600004C.

APR is supported by R01HL148565. MCH is supported by a grant from the National Heart, Lung, and Blood Institute (K08HL166687) and the American Heart Association (940166, 979465). PD reports funding from NCI (R01 CA248747, R01 CA260615) and DOD (W81XWH-22-1-0375). AGB has received support from a Burroughs Wellcome Foundation Career Award for Medical Scientists and the NIH Director’s Early Independence Award (DP5-OD029586). PN is supported by grants from the National Heart, Lung, and Blood Institute (R01HL142711, R01HL148050, R01HL148565) and Fondation Leducq (TNE-18CVD04).

## Disclosures

MCH reports consulting fees from CRISPR Therapeutics, advisory board service for Miga Health, and research support from Genentech. PN has received grant support from Amgen, Apple, AstraZeneca, Boston Scientific, and Novartis; spousal employment and equity at Vertex; has received consulting income from Apple, AstraZeneca, Novartis, Genentech/Roche, Blackstone Edwards Lifesciences, Foresite Labs, and TenSixteen Bio; and has served on the scientific advisory board of and holds equity in TenSixteen Bio and geneXwell, all unrelated to this work. AGB is the founding scientific advisor to and a shareholder of TenSixteen Bio. PD has received research funding from Janssen Research. The remaining authors have nothing to disclose.

## Notes

### Author Declarations

The Institutional Review Board at the Fred Hutchinson Cancer Center approved the studies and participants provided written informed consent.

## References

1. Jaiswal S, Natarajan P, Silver AJ, et al. Clonal Hematopoiesis and Risk of Atherosclerotic Cardiovascular Disease. N Engl J Med. 2017;377(2):111–121.

2. Yura Y, Sano S, Walsh K. Clonal Hematopoiesis: A New Step Linking Inflammation to Heart Failure. JACC Basic Transl Sci. 2020;5(2):196–207.

3. Yu B, Roberts MB, Raffield LM, et al. Supplemental Association of Clonal Hematopoiesis With Incident Heart Failure. J Am Coll Cardiol. 2021;78(1):42–52.

4. Schiattarella GG, Sequeira V, Ameri P. Distinctive patterns of inflammation across the heart failure syndrome. Heart Fail Rev. 2021;26(6):1333–1344.

5. Svensson EC, Madar A, Campbell CD, et al. TET2-Driven Clonal Hematopoiesis and Response to Canakinumab: An Exploratory Analysis of the CANTOS Randomized Clinical Trial. JAMA Cardiol. 2022;7(5):521–528.

